# The effect of leisure activities on quality-of-life scores for children with complex needs: A service evaluation in Wales, UK

**DOI:** 10.1101/2023.12.22.23300435

**Authors:** Nicole McGrath, Fiona Astill, Bethan Collins, Sabine Maguire, Alison Kemp, Lisa Hurt

**Author notes:** Corresponding author* Sabine Maguire, School of Medicine, Cardiff University, University Hospital of Wales, Cardiff CF14 4YS.

## Abstract

**Purpose:** Recent guidance has resulted in an increased level of interest in the wellbeing of children and young people, including those with complex needs. Evaluation of quality of life in this population is notoriously difficult, but has become increasingly vital when assessing the value of a service.

**Methods:** A previously validated tool, Quality of life Inventory-Disability (QI-Disability), was used in conjunction with parental reports on quality-of-life measures for children and young people before and after 6 and 12 months of attending specialist leisure activities provided by a charity (Sparkle) at children’s centres in South Wales.

**Results:** QI-Disability scores improved overall after 6 and 12 months of attending Sparkle club activities. However, the only statistically significant improvement was in the QI-Disability positive emotions domain. Parental reports also confirmed that children and young people were making progress towards their personal goals.

**Conclusion:** Collecting evaluation data within real-world services is challenging but essential. This paper uses quality-of-life measures to demonstrate how leisure activities provided by Sparkle improve scores for children with disabilities, including evidence of the perceived value for children, young people and parents.

**Plain English summary:** There is little previous research evaluating specialist leisure activities provided for children and young people with disabilities within real-world services. This evaluation aimed to find out if specialist leisure activities improved wellbeing scores for children with complex needs. We used parent-report questionnaires to measure changes to children’s quality of life whilst accessing these leisure activities. We found there is a benefit to children and young people with complex needs and their families when they participate in specialist leisure activities, and children experienced more positive emotions after accessing for 6 months.

## Introduction

Sparkle (South Wales) is the charity partner of Serennu, Nevill Hall and Caerphilly Children’s Centres, supporting children and young people (CYP), aged 0-17, with a range of disabilities and developmental difficulties, and their families. From the Centres and other venues in Gwent, South Wales, UK, Sparkle delivers specialist leisure activities for CYP with complex needs, including weekly play and youth clubs. The aim of this provision is to provide these CYP with equitable leisure opportunities to neurotypical and non-disabled peers, while supporting them to develop social skills and independence.

The needs of CYP with complex disabilities has recently been highlighted in updated National Institute for Health and Care Excellence (NICE) guidance within the UK [1]. NICE have defined this group as CYP (aged 0-25 years) who require coordinated education, health and social care support because of severe and complex needs, and who are eligible for education, health and care plans (or individual development plan in Wales). The total number of CYP with complex needs in the UK continues to rise [2]; these children are likely to require additional support from health, education and social care services at various points during their childhood.

The NICE guidance includes greater emphasis on improving *wellbeing* in this population, as well as the development of essential skills for adulthood, which are often overlooked. This includes the benefits of participation in social activities. The recommendations recognise the current shift towards integration for CYP with complex needs. However, while there are notable advantages for children of all abilities participating in activities together, evidence also indicates that CYP with complex needs have additional barriers to participation in ‘inclusive’ leisure activities [1]. In an era of extreme cost pressures, it is vital to ensure that leisure services designed specifically for these young people’s needs are adding value to their lives.

One key outcome in such an evaluation is quality of life (QoL). QoL is described by the World Health Organisation as an individual’s perceptions of their life *“in the context of the culture and value systems in which they live and in relation to their goals, expectations, standards and concerns”* [3]. Extensive research into how the health of an individual effects their QoL (termed health-related QoL) has highlighted how health issues can affect an individual’s interactions with others and societal perceptions. Previous research has indicated that CYP with complex needs associated with intellectual disabilities and/or social communication difficulties experience poorer QoL, which has in part been linked to limitations in community activities [4]. Evidence also suggests that activities which enable social inclusion for CYP with physical disabilities, are linked to improved mental health outcomes [5].

It has been shown that childhood disability impacts on the QoL for the entire family. Leung and Li-Tsang [6] demonstrated that parental QoL scores were correlated with severity of a child’s disability; the most prominent effect was seen in the domains of physical and psychological health, and environment when their children had severe functional impairment. Parents of children with disabilities are more likely to limit their social activities due to anxiety about their child’s behaviour in a public place, and they may have reduced socialising with family and friends, partly due to the time taken attending appointments or caring for their children [7]. A lower QoL score for parents in the environment domain was highly correlated to the severity of the child’s disability, and reflects the limited opportunities they have to manage or schedule activities for themselves [6]. Davies and Gavidia-Payne [8] assessed the factors that impact QoL in siblings of CYP with complex needs. In this study, support from trusted professionals was found to have a positive impact on quality of family life, suggesting that interventions that improve QoL for disabled children *could* have a significant positive impact on other members of the family, enhancing the value added by these services.

Sparkle leisure activities consist of 1–2-hour clubs once per week where CYP can attend without needing parental support (Figure 1). The sessions have a minimum ratio of staff to participants, which vary depending on the needs of the CYP in each club, but can include 1:1 and 2:1 support. One important factor enabling participation for CYP in Sparkle clubs is that the Leisure Support Workers are trained to support those with complex needs, including support for medical needs (such as first-aid, medication administration and epilepsy training) and communication needs (including sign language and autism awareness). Another key feature for these leisure clubs is the child-led focus; CYP are given a choice of activities (rather than this being dictated by the organisers), and efforts are made to ensure that there is sufficient variety in the activities on offer between sessions, ensuring CYP become accustomed to changes as part of their overall social development and allowing them to explore personal interests.

**Figure 1:**
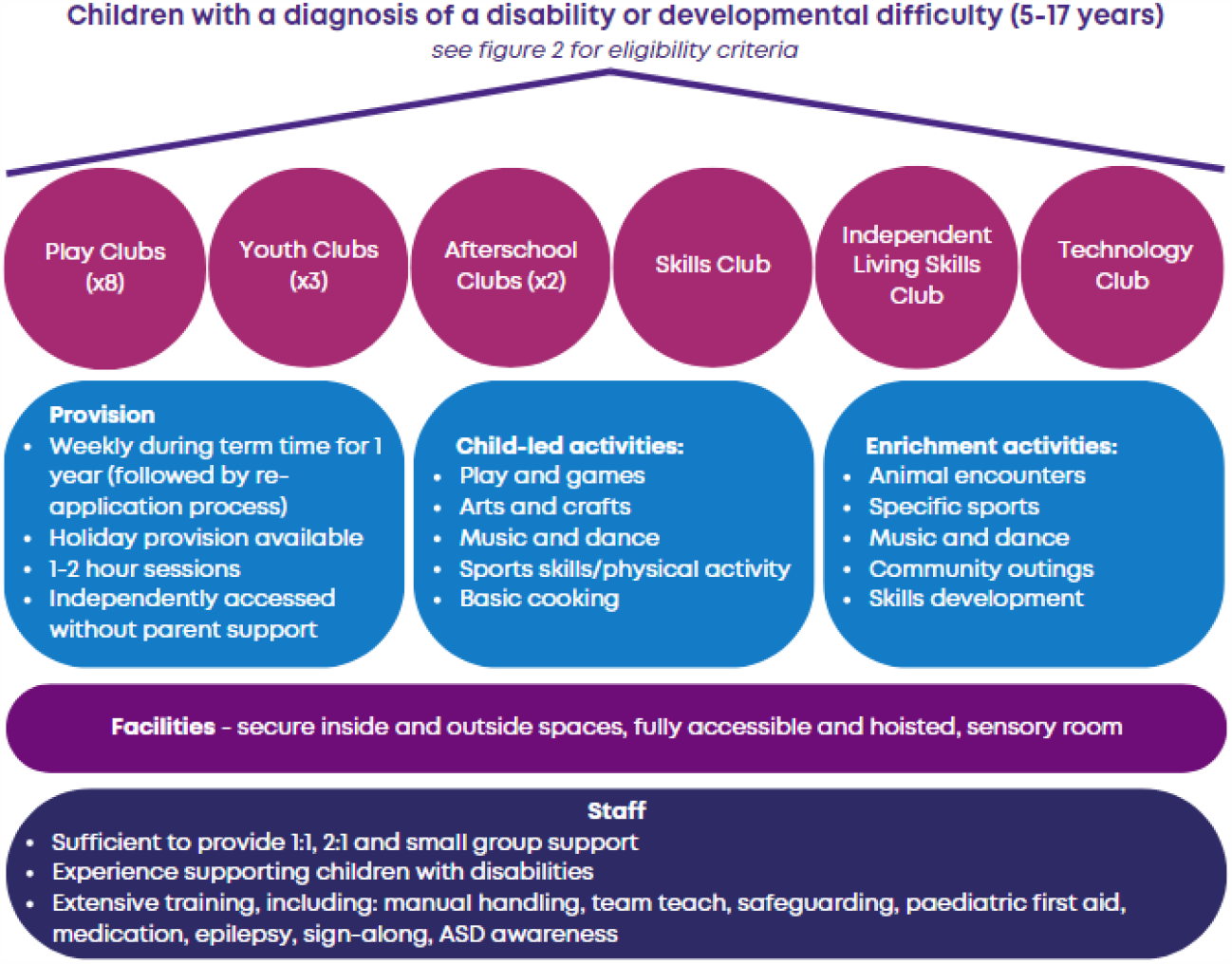
Specialist leisure provision for children aged 5-17 years with complex disabilities, provided by Sparkle (South Wales).

The aim of this study was to evaluate the impact of Sparkle’s specialist leisure provision on QoL for CYP with disabilities and developmental difficulties, using a previously validated tool aimed at carers of children with disability, the Quality-of-Life Inventory-Disability (QI-Disability). This has been completed as part of a larger service evaluation of Sparkle’s leisure activities using quantitative and qualitative methodologies.

## Method

### Design

This is a prospective quantitative descriptive case series conducted over a one-year period.

### Setting

We recruited participants attending Sparkle club activities from sites across South Wales. These clubs cater for CYP with a range of disabilities and developmental difficulties, including Autistic Spectrum Disorder, Attention Deficit & Hyperactivity Disorder, syndromes, cerebral palsy, or combinations of the above. CYP are referred to Sparkle club services by their caregivers, and require confirmation of their complex needs by either a medical or social care professional involved in their care (for eligibility criteria, see Figure 2). The specific needs of each participant are assessed prior to joining and the structure of each session can be adapted to the needs of the attendees (see appendix 1). Places in the clubs are allocated on a needs basis, and on average 52% of children attending require 1:1 or 2:1 support due to needs such as: non-verbal communication; unpredictable behaviour putting themselves or others at risk; and/or severe physical disability (e.g., spastic quadriplegia). The activities on offer during Sparkle leisure clubs vary (figure 1), but CYP are facilitated to choose which activities that they wish to participate in.

**Figure 2:**
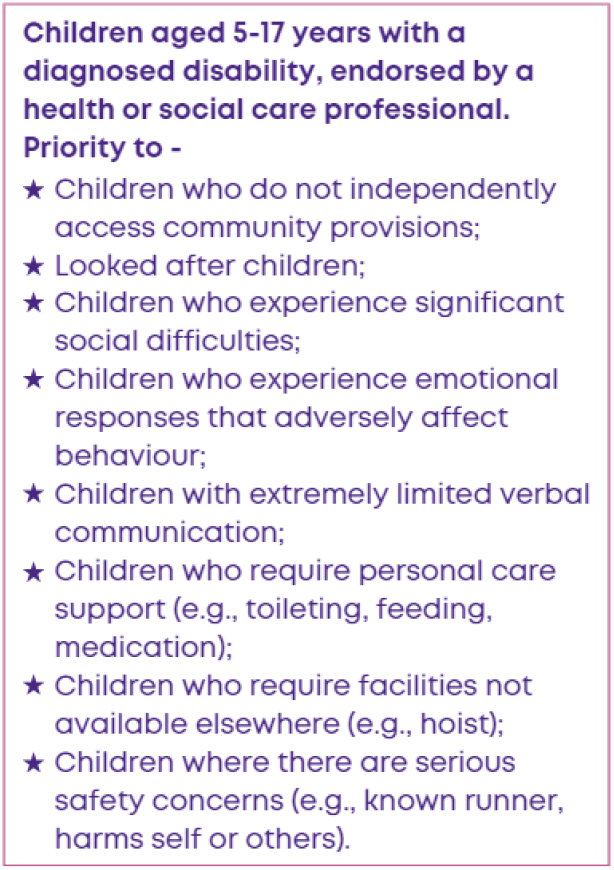
Eligibility criteria for Sparkle (South Wales)’s specialist leisure provision for children aged 5-17 years with complex disabilities.

### Participants

Between November 2020 and September 2022, all new referrals to Sparkle’s leisure activities were invited to take part in the evaluation. Due to the high demand for Sparkle leisure services, it is usual for children to be on a waiting list for a place in club for some weeks, depending on the availability of staff and/or funding. All children meeting the Sparkle eligibility criteria for clubs were eligible for inclusion in this evaluation.

Of the 148 parents/carers invited to participate in this evaluation, 87 agreed to take part, all of whom completed the baseline QI-Disability questionnaire (Figure 3). At the end of data collection, 29 of these children had not been attending club for 6 months (with the varying start time in club due to time spent on the waiting list), and could not therefore be included in the analysis. Of the remaining 58 parents/carers, 38 completed both baseline and 6 month questionnaires; 13 of these also completed 12 month questionnaires at the end of data collection period (September 2022).

**Figure 3:**
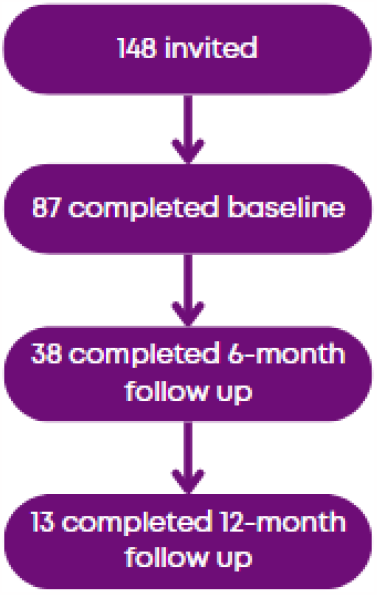
Flowchart of parent/carer participation rates in an evaluation of the impact of specialist leisure provision for children and young people with complex needs.

### Ethical approval

The service evaluation was approved by Aneurin Bevan University Health Board Research and Development Research Risk review panel on 30^th^ June 2020. Parents were given written information about the evaluation, and provided consent for their children to be included. Participants were assured that their participation (or not) did not have any effect their subsequent offer of leisure services. Data was pseudo-anonymised at the point of collection. All data collected was stored securely; physical data were stored securely on-site, and digital data remained within the local health board intranet system.

### Data collection

The QI-Disability is a parent-reported measure of QoL for children aged 5-18 years with intellectual disabilities. It was designed in collaboration with families and carers of CYP with intellectual disabilities and measures QoL across six domains; physical health, positive emotions, negative emotions, social interactions, leisure and outdoors, and independence. The QI-disability uses 36 questions in total [9], each rated on a Likert scale from never (score 1) to very often (score 5) with reverse scoring for the negative emotions domain. Scores are then transformed; 1 to 0, 2 to 25, 3 to 50, 4 to 75, and 5 to 100. The questionnaires were distributed to parents/carers who consented to participate via email and completed online. Paper questionnaires were also available for parent/carers requesting them. Parents/carers completed the questionnaire whilst on the waiting list for the clubs (baseline), and subsequent questionnaires when their child had been accessing Sparkle activities for 6 months and 12 months. The scores for individual participants were collated at each time point and analysed for any change over time.

In addition to the QI-Disability questionaries, parents/carers were asked to identify specific goals for their child whilst attending leisure clubs during their profile interview when they first joined Sparkle. At 6 and 12 months, parents/carers were asked to reflect on their child’s progress against these anticipated goals and whether they and their family had experienced any impact on their overall wellbeing since accessing (Table 1).

**Table 1:**
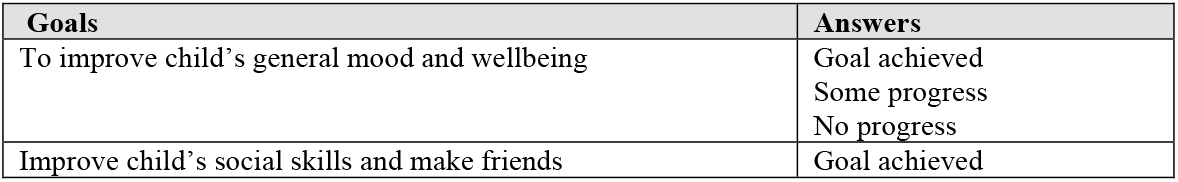
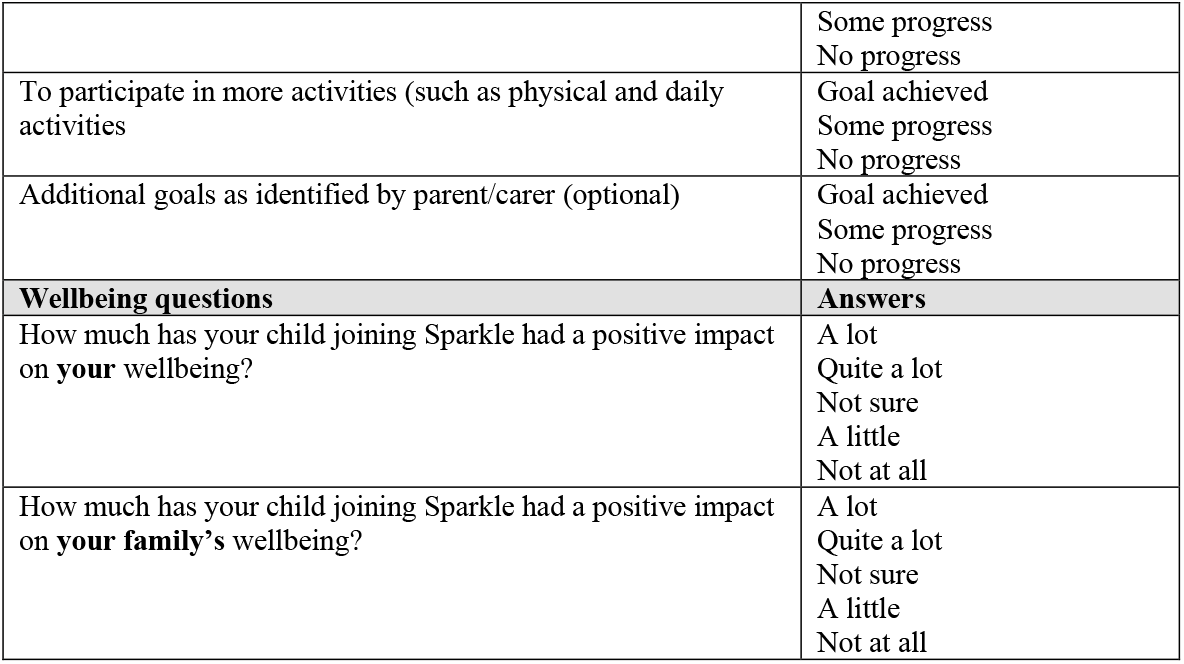
Assessment of progress against goals identified by parent/carers prior to their child commencing specialist leisure provision and additional information on the impact of the club on the parent/carer and family, assessed after 6 months and 12 months of attendance at Sparkle leisure activities.

| Goals | Answers |
| --- | --- |
| To improve child's general mood and wellbeing | Goal achieved<br>Some progress<br>No progress |
| Improve child's social skills and make friends | Goal achieved |
|  | Some progress<br>No progress |
| To participate in more activities (such as physical and daily activities) | Goal achieved<br>Some progress<br>No progress |
| Additional goals as identified by parent/carer (optional) | Goal achieved<br>Some progress<br>No progress |
| **Wellbeing questions** | **Answers** |
| How much has your child joining Sparkle had a positive impact on **your** wellbeing? | A lot<br>Quite a lot<br>Not sure<br>A little<br>Not at all |
| How much has your child joining Sparkle had a positive impact on **your family's** wellbeing? | A lot<br>Quite a lot<br>Not sure<br>A little<br>Not at all |

### Analysis

This is a descriptive analysis of the quantitative data collected during this service evaluation, and a summary of children’s achievement against the pre-set goals identified by parents/carers. Mean scores for the QI-Disability at the different time points were examined and compared using a Wilcoxon signed rank statistical test. The percentage of children making progress against their pre-set goals was also calculated, with responses of ‘goal achieved’ and ‘some progress’ considered as a positive response for the goal questions; ‘a lot’ and ‘quite a lot’ of improvement was considered as a positive response for the family impact questions.

## Results

### Demographics

Of the 38 children with baseline and 6 month follow-up data, 34 were male and 4 were female. The average age was 9 years (median = 9, range = 12). The majority of participants (87%, n=33) had a diagnosis of neurodevelopmental disability, including autism, and 5 (13%) had a physical disability such as cerebral palsy and severe epilepsy; 9 CYP (24%) had severe communication difficulties (‘non-speaking’).

Further analysis of the 13 participants who provided data at 12 months was completed to ensure the demographics remained consistent. This group included 13 males (100%) with an average age of 9 years (median = 9, range = 8). In this group, 10 had a diagnosis of neurodevelopmental disability (77%) and 3 of physical disabilities (23%); 3 children (23%) were identified as non-speaking.

### QI-Disability

Figure 4 shows the QI-Disability scores at the different measurement time points. There was an upward trend for four domains (positive emotions, negative emotions, social interactions and independence) from the baseline questionnaires to the 12-months post-intervention questionnaires. Only the positive emotions domain provided a statistically significant difference; increasing from 72.35 at baseline to 78.22 after six months (Wilcoxon signed-rank test, *z* = -2.5618, *p* < .05.). The data also suggests an improvement in overall QoL scores after attending 6 months of Sparkle activities, but this comparison between baseline and 6 months was not statistically significant.

**Figure 4:**
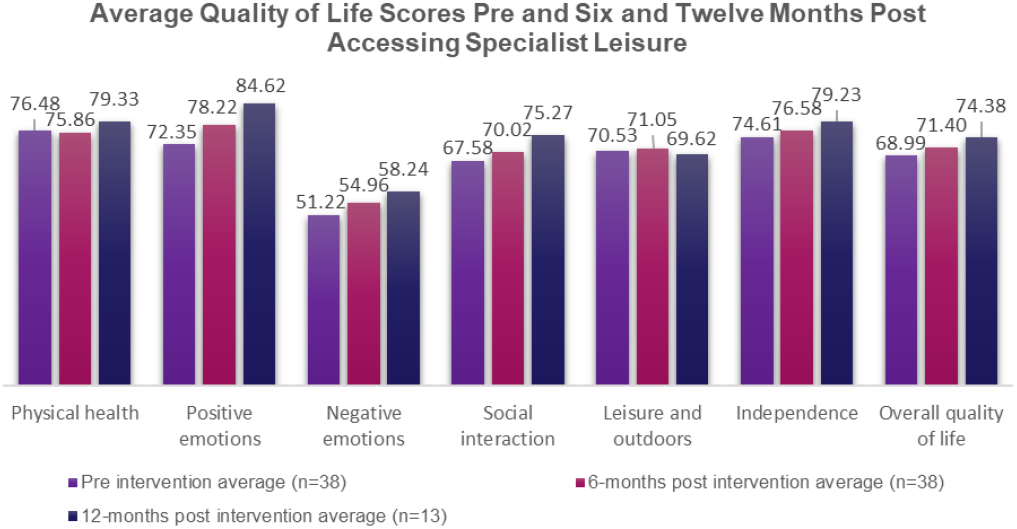
QI-Disability scores before accessing specialist leisure provision, and after 6 and 12 months of accessing the provision (the ‘negative emotions’ domain was reverse scored, therefore an increase in this score suggests a decrease in negative emotions experienced).

### Goals

A high proportion of parents/carers identified an improvement against personal goals for their child at 6 and 12 months; most achievement was made against goals relating to mood and wellbeing at 6 and 12 months, and participating in more activities at 12 months (Table 2).

**Table 2:** Percentage of children/young people who had made progress against/achieved goals as set by their parent/carers at baseline, reassessed at 6 and 12 months.

|  | 6 months (n=38) | 12 months (n=13) |
| --- | --- | --- |
| **Improve mood and general wellbeing** | 94% | 92% |
| **Develop social skills and make friends** | 89% | 77% |
| **Participate in more activities** | 87% | 92% |

### Family and parent/carer wellbeing

Many parents/carers (72%) felt their own wellbeing improved due to their child accessing Sparkle clubs for 6 months, rising to 85% following 12 months. Fewer felt there was an improvement in their wider family’s wellbeing – 66% at 6 months, and 72% at 12 months.

## Discussion

The results of this service evaluation indicated a positive impact on QoL scores for CYP participating in specialist leisure activities at 6 and 12 months, although only one domain of the QI-Disability (positive emotions) showed a statistically significant change. The additional questions regarding the goals set by parents/carers and family wellbeing also showed positive impacts.

The main aim of this work was to evaluate the effect of participation in specialised leisure activities on the QoL of CYP with complex needs and their families. The leisure services are funded through charitable donations and evaluation data is essential to assess which activities should continue and which are not providing value-for-money. In addition, given that the service has several *unique features* (CYP with wide ranging complexity catered for, with 1:1 or 2:1 support where necessary, highly trained leisure staff, child-led activities, tailored to cognitive ability, integrated enrichment, etc.), collecting data from this population was also considered important to inform the development of other similar services. Conducting evaluations on a ‘real world’ service such as this, which caters for a wide range of clinical conditions, is challenging. The use of a validated QoL tool enabled us to obtain quantitative data in a manner acceptable to parents/carers, and it allowed comparisons to be made between an otherwise highly heterogeneous group of individuals. It also allowed us to examine how the service affected key areas of *health and wellbeing* in this group, which are increasingly recognised by NICE as fundamental in overall care provision for this population. As such, this work contributes unique data relating to children with severe complex needs, who are an increasing population, currently missing from research data. Our work expanded on this by including questions linked to personal goals and the influence that a specialist leisure service has on family wellbeing, providing insight into the wider impact of leisure provision on the lives of these CYP. Wellbeing data is rarely collected by services such as these, despite this being a useful tool for demonstrating the value added by the service on both an individual and at a community level.

A particular challenge with service evaluations such as these is to recruit participants, especially for repeat evaluations, as is evident from the drop-off of parents/carers at the 6 and 12 month time points in this evaluation. The level of demand for Sparkle leisure services means that CYP are placed on waiting lists for variable lengths of time, resulting in some consenting families not having completed 6 months of attendance at the completion of the study. In addition, families of CYP with complex needs are likely to have limited time for participation in research activities, particularly while prioritising the best use of their ‘respite’ time while the CYP are attending leisure activities. To understand whether this was the case in this evaluation, informal conversations were instigated with parents/carers. These highlighted “data collection fatigue” in the parents/carers, given the multiple forms that they are asked to complete relating to their child’s health, education, financial support and so on. They also stated that, because of the time pressures associated with caring, many felt that it was not necessary for “*them*” to complete the questionnaires as “*others*” would have done so. No issues that may have biased the responses of the parents/carers about the effects of the activities over time were identified during these conversations (such as a dissatisfaction with the activities). The drop-off in completion of the QI-Disability scores at 6 and 12 months has therefore limited the power of the analysis to identify changes over time, but we do not believe that the group who responded were a biased sample. The follow-up conversations have also allowed the team to identify key messages to provide to parents/carers to improve response rates in future data collection rounds, such as how essential evaluative data is when seeking grant funding for the services.

The individual CYP recruited to this study had both neurodevelopmental disorders and/or neuro and physical disabilities, with variable levels of communication difficulties. Available validated QoL assessment tools for this population are limited, and whilst the physical health domain may be expected to vary for CYP with a physical disability, this domain may be more stable for those with neurodevelopment disorders, with the latter being more challenged by the emotional and social domains. Thus it is unsurprising that no significant changes were seen in the physical health or ‘independence’ elements of the QI-Disability tool.

This evaluation was conducted in a ‘real world’ setting. As such, the content of the specific clubs that children attended was not standardised, as it may be in a research study. However, all clubs were run using the same principles of ensuring that every child, regardless of the complexity of their disability, should have an opportunity to engage in rewarding activities, socialise with others, and experience opportunities they would not otherwise have (e.g., participating in sports, music, and so on). Furthermore, the variation between club activities is a deliberate feature of the club’s child-led design. The choice of leisure activities available to the CYP may be modified by the Sparkle club organisers (to avoid CYP participating in a single preferred activity during every session), but this may lead to some CYP participating in a greater proportion of activities that develop a specific skill set. There is not therefore a standard “intervention” being tested in this evaluation and it is not possible to examine whether specific activities are particularly beneficial to the CYP. However, empowering CYP with disability via the child-led ethos, and the range of experiences that this provides, is a key feature of this service model, regardless of the challenges that this poses to evaluation. It is clear from the significant improvements made by CYP against the goals set for them by their parents/carers at the outset, that this service is meeting their needs and is enhancing their QoL, with the added benefit that this has on the children’s home life and parental wellbeing.

The *challenges* in collecting data from CYP with complex needs has resulted in a dearth of information in this area, despite the need to understand whether the services provided for them are effective; in fact, many researchers omit children with the most complex needs from their studies entirely [10]. Previous research has rarely attempted to include this population of CYP when assessing services, most often relying on the views of parents as a proxy, such as in this evaluation. People familiar to the CYP, who are responsive to their needs and care about them, are thought to be the best proxy respondents [11]. Numerous tools have been produced to attempt to measure CYP’s QoL, with or without parent involvement [12]; however, it may be that theses tools are not suitable for a population of children with complex disabilities [13]. Previous studies have shown significant differences between a child’s score and a mother’s opinion score [14], therefore demonstrating that parents may not always be able to give an accurate reflection of their child’s QoL. Previous research that has attempted to include feedback from CYP with communication difficulties has demonstrated the role of qualitative data collection [15], and case study work with population, including the direct views of parents/carers, leisure staff and the CYP themselves, found positive psychosocial outcomes linked to leisure participation [16].

### Further areas of research

The difficulties of obtaining meaningful feedback from CYP with communication difficulties has been highlighted as a significant limitation in this service evaluation. This is an important area of ongoing research, which is unlikely to have one overall solution due to the heterogeneity of CYP with disabilities. However, this evaluation has demonstrated the positive impact specialist leisure provision can have for CYP with disabilities and their families, and it is recommended that future research should explore the impact of specialist leisure for this group in more detail, including the direct views of the CYP themselves, to ensure the best possible outcomes for CYP with complex needs and influence service delivery in other areas.

## Conclusion

It has been highlighted by NICE something that parents/carers have always known, namely that determining the wellbeing of children with complex needs is as important as their physical/medical needs. This requires leisure services offering them the same experiences and opportunities as their neurotypical or non-disabled peers, while meeting their complex needs. This study provides evidence of the positive effects on the QoL of CYP with disabilities who attend specialist leisure provision, in this case by Sparkle, a charity. In addition, the analysis supports the link between this service and improved wellbeing for parents/carers and their families. Future work is ongoing to facilitate collection of direct feedback from this population of CYP.

## Data Availability

Data produced in the present study are not openly available due to lack of participant consent to open the data.

## Acknowledgements

The authors would like to thank the parents and carers who participated in this evaluation.

## Statements and Declarations

### Funding

No funding was received to conduct this evaluation.

### Competing interests

One of the authors is employed and funded by Sparkle (South Wales), the registered charity (1093690) that delivers the service evaluated during this research. Another author is an unsalaried Trustee of the charity. The authors have no other conflicts of interest to declare.

### Author contributions

All authors contributed to the study conception and design. Material preparation, data collection and analysis were completed by Bethan Collins, Fiona Astill and Nicole McGrath. The first draft on the manuscript was written by Nicole McGrath and Bethan Collins, and all authors commented on and/or edited various versions of the manuscript. All authors read and approved the final manuscript.

### Ethics approval

The service evaluation was approved by Aneurin Bevan University Health Board Research and Development Research Risk Review panel on 30^th^ June 2020.

### Consent to participate

Informed, written consent was obtained from all participants in this evaluation. Participants were assured that their participation (or not) did not have any effect their subsequent offer of leisure services.

### Consent to publish

Participants gave consent for anonymised group data to be published following the evaluation.

## Appendix 1: Aims of specialist leisure clubs delivered by Sparkle (South Wales)

| **Club** | **Aims of clubs** |
| --- | --- |
| Youth Clubs | Independence, social skills, communication skills, interacting with others/<br>building relationships |
| Play Clubs | Play opportunities, children engaging with resources, social skills,<br>communication, friendships |
| After School Club (5-11 years) | Play opportunities, children engaging with resources, social skills,<br>communication, friendships |
| After School Club (12-17 years) | Independence, social skills, communication skills, interacting with others/<br>building relationships |
| Skills Club | Physically active, building stamina, co-ordination and social skills, team work |
| Independent Living Skills | Independence, skills for independent living |
| Technology Club | Develop skills in technology, interaction with peers, communication skills, turn<br>taking, managing frustrations (gaming related) |

## Notes

### Author Declarations

The Research Risk Review Panel of Aneurin Bevan University Health Board Research and Development Department gave ethical approval for this work.

